# Changes in sexual behavior, PrEP adherence, and access to sexual health services due to the COVID-19 pandemic among a cohort of PrEP-using MSM in the South

**DOI:** 10.1101/2020.11.09.20228494

**Authors:** Sanjana Pampati, Kayla Emrick, Aaron J. Siegler, Jeb Jones

## Abstract

**Background:** The COVID-19 pandemic has had unforeseen consequences on the delivery of HIV and STD prevention services. However, little is known about how the pandemic has impacted PrEP-using men who have sex with men (MSM).

**Methods:** Data come from an online cohort of PrEP-using MSM in the Southern United States from October 2019 to July 2020. Participants were administered ten surveys in total, including one ad hoc survey specifically on COVID-19. We conducted a cross-sectional analysis of this ad hoc survey (n=56) and present changes in sexual behaviors and utilization of and access to sexual health services. Using linear mixed-effects regression models, we also analyzed data from the larger cohort (n=78) and document how sexual behaviors and PrEP use varied longitudinally focusing on three months: February (pre-pandemic), April (early pandemic), and June (later in the pandemic).

**Results:** A fifth of participants discontinued or changed how often they take PrEP because of COVID-19. A quarter of the cohort documented challenges when attempting to access PrEP, HIV testing, or STD testing. For all sexual behaviors examined longitudinally—number of male sexual partners, anal sex acts, condomless anal sex, oral sex (all measured in the past 2 weeks)—there was a significant decrease from February to April followed by a significant increase from April to June.

**Discussion:** Our findings suggest reduced access to and utilization of STD and HIV services coupled with a continuation of behaviors which confer STD/HIV risk. Ensuring appropriate delivery of STD/HIV prevention services during this pandemic is imperative.

## Introduction

As of November 2020, there were almost 10 million confirmed coronavirus disease 2019 (COVID-19) cases and over 230,000 deaths across the United States.^1^ Efforts to control the pandemic have heavily relied on the implementation of social distancing guidelines^2^ and stay-at-home orders^3^ that intend to limit the number of social contacts and promote isolation at home. Such practices can have unforeseen consequences on the delivery of needed health services.

Evidence suggests the pandemic has already influenced access to and utilization of STD/HIV services, including a decline in HIV post-exposure prophylaxis prescriptions,^4^ potential disruptions in the HIV care continuum,^5-7^ and access to HIV/STD testing.^8^ In the context of delivery of sexual health services, MSM who use HIV pre-exposure prophylaxis (PrEP) in the South are a key population of interest, as prescription guidelines for PrEP underscore the importance of routine health care visits and quarterly HIV and STD testing for MSM at high risk for recurrent STDs.^9^ Further, the focus on PrEP users in the Southern United States is warranted; the burden of HIV and STDs is disproportionately concentrated in the South^10,11^ and many of the current COVID-19 hotspots are in the South.^1^ Refills for PrEP are typically authorized at a check-up visit after negative HIV status is confirmed and prescriptions may require pick-up at a pharmacy. As a result, it is possible that access and adherence to PrEP may be affected by the COVID-19 pandemic, as well as compliance to routine HIV/STD testing.

Elucidating changes in sexual behaviors due to the pandemic is also important. In fact, reduced access to STD/HIV testing and PrEP discontinuation may be less concerning if coupled with a reduction in sexual risk. In the earlier stages of the pandemic, a nationwide study with MSM suggested reductions in sexual risk taking, including decreases in the number of sexual partners.^5^ However, with the ongoing lifting of stay-at-home orders and declining compliance to social distancing guidelines,^12^ it is not apparent if these decreases in sexual risk have persisted. There is a need to examine longitudinal trends in sexual behaviors in the context of the ongoing pandemic. Moreover, little is known about individuals’ COVID-19 risk perceptions in the context of sexual behavior and such data can help contextualize behavioral changes. Accordingly, we examine changes to access and utilization of sexual health services and changes in sexual behavior due to the COVID-19 pandemic, as well as COVID-19 risk perceptions among a cohort of PrEP-using MSM in the South.

## Methods

### Data collection

Data come from a cohort study examining trajectories of PrEP use among MSM recruited online in the Southern United States from October 2019 to July 2020. Eligible participants were cisgender male, 18-34 years old, lived in the Southern United States and planned to stay there for at least 16 weeks, English proficient, reported anal sex with a man in the past 6 months, were HIV-negative, and were current users of oral HIV PrEP. Participants were administered a baseline survey, seven identical biweekly surveys assessing sexual behavior and PrEP use, and a final survey on sexual behavior and future plans for PrEP use. As the end of the study’s timeline coincided with the COVID-19 pandemic, an ad hoc survey assessing how the pandemic has influenced participants’ sexual behavior, PrEP use, and access to sexual health services was administered in June-July 2020 to all currently and previously enrolled participants, and will hereafter be referred to as the COVID-19 survey. Participants were compensated for their participation.

### Measures

The COVID-19 survey included questions assessing how the pandemic and efforts to control it have influenced access to and utilization of sexual health services (e.g., STD testing, HIV testing, PrEP) and changes relating to a spectrum of sexual activities (e.g., anal sex, sexual activity with causal partners). Participants were asked to indicate the perceived level of COVID-19 acquisition risk on a scale of 0 (no risk) to 100 (highest risk) associated with specific sexual behaviors. Additionally, each of the seven check-in surveys, the final survey, and the COVID-19 survey included identical items measuring in the past 2 weeks: number of missed PrEP doses, number of male sexual partners, anal sex acts, condomless anal sex, and oral sex.

### Analysis

This study includes both cross-sectional and longitudinal components. First, we conducted a cross-sectional analysis of the COVID-19 survey (n=56), presenting percentages and number of respondents endorsing response options for questionnaire items. We computed the median and interquartile range for the four items assessing perceived risk of COVID-19 acquisition for specific sexual behaviors.

For longitudinal analyses, we utilized data from the larger cohort study (n=78) and fit linear mixed-effects regression models to examine potential period effects for the number of missed PrEP doses, number of male sexual partners, any anal sex with male partners, any condomless anal sex with male partners, and any oral sex with male partners, all measured in the past 2 weeks. The calendar month the survey was submitted was the independent variable. Using these models, means/proportions and standard errors of each repeated measure during three months [February (pre-pandemic), April (early pandemic), and June (later in the pandemic)] were computed. T-tests were used to examine differences by month, and findings were considered statistically significant if p<.05. All analyses were conducted in SAS 9.4 (SAS Institute, Cary, NC).

## Results

Among those who completed the COVID-19 survey (n=56), the mean age was 26.3 years (standard deviation = 4.2) with a range of 19 to 34. The sample was 64% white (n=36), 18% Black (n=10), and 14% Hispanic (n=8). Table 1 presents changes in STD/HIV testing, PrEP use, and sexual behaviors from the COVID-19 survey. Five participants (9%) reported discontinuing PrEP use. Several participants reported difficulties obtaining their PrEP medication (n=8, 16%). Many participants did not have an HIV test (n=18, 32%) or STD test (n=24, 43%) in the past 3 months. Further, several cited difficulty obtaining an HIV test (n=11, 20%) or STD test (n=10, 18%) due to the pandemic.

**Table 1.** Changes in PrEP use, STD/HIV testing, and sexual behavior due to COVID-19 pandemic among PrEP-using MSM (n=56)

| **Experience** | **n (%)** |
| --- | --- |
| Change in PrEP adherence |  |
| Taking PrEP less frequently | 6 (11) |
| Discontinued PrEP use | 5 (9) |
| No change | 45 (80) |
| Difficulties obtaining PrEP medication<sup>1</sup> |  |
| Yes | 8 (16) |
| No/I haven't tried to get my PrEP medication | 42 (84) |
| Change in how PrEP is obtained<sup>1</sup> |  |
| Yes | 7 (14) |
| No | 43 (86) |
| Last HIV test |  |
| Within past 3 months | 38 (68) |
| 4-6 months ago | 15 (27) |
| 7-12 months ago | 3 (5) |
| Last STD test |  |
| Within past 3 months | 32 (57) |
| 4-6 months ago | 18 (32) |
| 7-12 months ago | 4 (7) |
| >1 year ago | 2 (4) |
| Difficulty obtaining HIV test |  |
| Yes | 11 (20) |
| No | 34 (61) |
| Have not tried | 11 (20) |
| Difficulty obtaining STD test |  |
| Yes | 10 (18) |
| No | 31 (55) |
| Have not tried | 15 (27) |
| Had sex since social distancing guidelines began | 44 (79) |
| Sex with someone who lives with me<sup>1</sup> | 3 (7) |
| Sex with someone who does not live with me that lives alone<sup>1</sup> | 33 (75) |
| Sex with someone who does not live with me that lives with other people<sup>1</sup> | 25 (57) |
| Sex with someone who does not live with me whose living situation is unknown<sup>1</sup> | 4 (9) |
| Sexual activity with casual partners |  |
| Decreased | 46 (82) |
| No change | 7 (12) |
| Increased | 1 (2) |
| Does not apply | 2 (4) |
| Sexual activity with main partner |  |
| Decreased | 21 (38) |
| No change | 17 (30) |
| Increased | 7 (12) |
| Does not apply | 11 (20) |
| Frequency of kissing |  |
| Decreased | 34 (61) |
| No change | 18 (32) |
| Increased | 2 (4) |
| Does not apply | 2 (4) |
| Frequency of oral sex |  |
| Decreased | 36 (64) |
| No change | 18 (32) |
| Increased | 1 (2) |
| Does not apply | 1 (2) |
| Frequency of anal sex |  |
| Decreased | 38 (68) |
| No change | 17 (30) |
| Increased | 1 (2) |
| Frequency of rimming |  |
| Decreased | 37 (67) |
| No change | 15 (27) |
| Increased | 1 (2) |
| Does not apply | 2 (4) |
| Number of sexual partners |  |
| Decreased | 44 (78) |
| No change | 12 (21) |
| Increased | 0 (0) |

<sup>1</sup>Asked among those who are currently taking PrEP
<sup>2</sup>Asked among those who reported having sex since the beginning of social distancing guidelines
Note: Percentages may not add to a 100% due to rounding.

Since the beginning of social distancing guidelines, 79% of participants reported having sex (n=44), including with someone who lives with them (n=3, 7%), someone who does not live with them that lives alone (n=33, 75%), and someone who does not live with them that lives with other people (n=25, 57%). The majority of participants reported decreases in all sexual behaviors examined, ranging from kissing (n=34, 61%) to anal sex (n=38, 68%) since the beginning of the COVID-19 pandemic. Additionally, most reported decreases in sexual activity with casual partners (n=46, 82%). In contrast, fewer participants reported decreases in sexual activity with a main partner (n=21, 38%).

Examining longitudinal trends in sexual behaviors (Table 2), the proportion reporting having ≥2 male sexual partner in the past 2 weeks was 0.32 in February (SE = 0.05), and this proportion significantly declined from February to April (proportion = 0.10, SE = 0.04), and then significantly increased from April to June (proportion = 0.26, SE = 0.06). The three other sexual behavior indicators – proportion having any anal sex, any oral sex, and any condomless sex—all measured in the past 2 weeks and with male partners also showed significant decreases from February to April and then significant increases from April to June. In contrast, there did not appear to be substantial variation in the mean number of missed PrEP doses by calendar month among those who remained on PrEP.

**Table 2.**
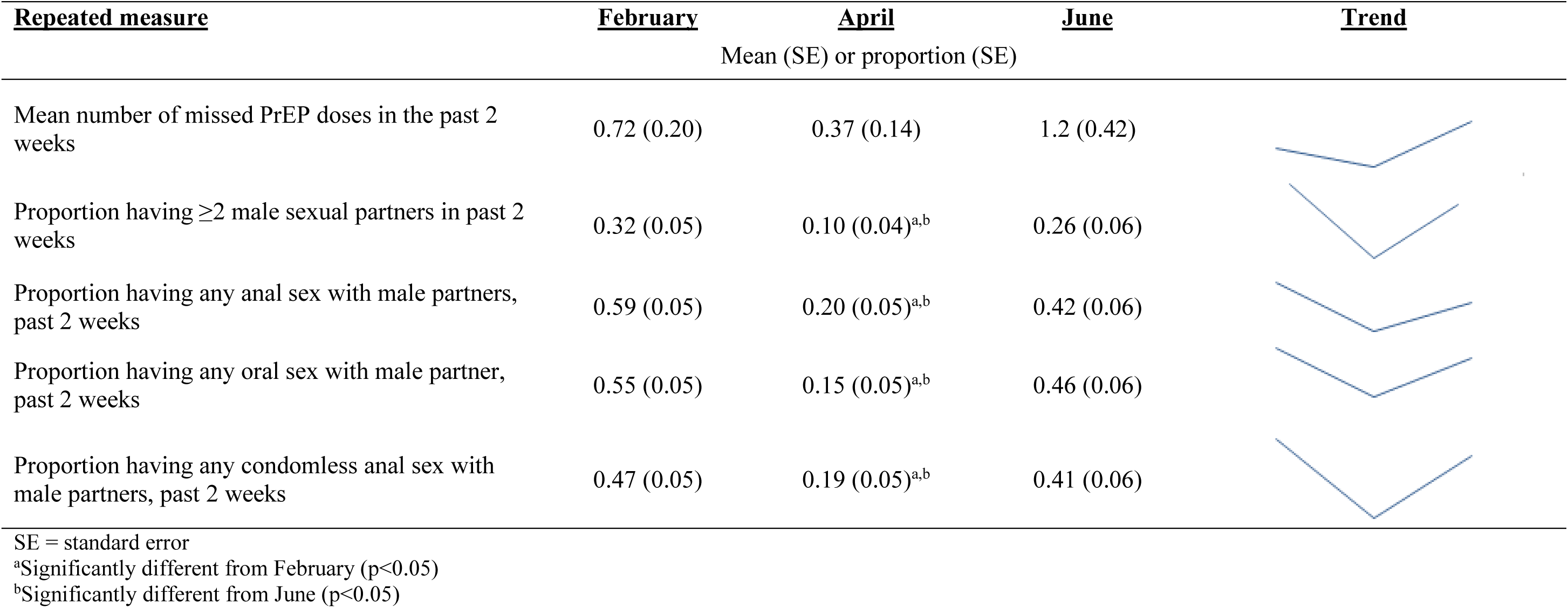
Longitudinal trends in PrEP use and sexual behaviors, February-June 2020

| **<u>Repeated measure</u>** | **<u>February</u>** | **<u>April</u>** | **<u>June</u>** | **<u>Trend</u>** |
| --- | --- | --- | --- | --- |
|  | Mean (SE) or proportion (SE) |  |  |  |
| Mean number of missed PrEP doses in the past 2 weeks | 0.72 (0.20) | 0.37 (0.14) | 1.2 (0.42) |  |
| Proportion having $\geq 2$ male sexual partners in past 2 weeks | 0.32 (0.05) | 0.10 (0.04)<sup>a,b</sup> | 0.26 (0.06) | |
| Proportion having any anal sex with male partners, past 2 weeks | 0.59 (0.05) | 0.20 (0.05)<sup>a,b</sup> | 0.42 (0.06) |  |
| Proportion having any oral sex with male partner, past 2 weeks | 0.55 (0.05) | 0.15 (0.05)<sup>a,b</sup> | 0.46 (0.06) |  |
| Proportion having any condomless anal sex with male partners, past 2 weeks | 0.47 (0.05) | 0.19 (0.05)<sup>a,b</sup> | 0.41 (0.06) |  |

SE = standard error
<sup>a</sup>Significantly different from February (p<0.05)<sup>b</sup>Significantly different from June (p<0.05)

On a scale of 0 (no risk) to 100 (highest risk), the median perceived level of COVID-19 acquisition risk was 100 (IQR: 14) for kissing; 74 for rimming (IQR: 66); 75 for oral sex (IQR: 67); 66 for anal sex (IQR: 75; data not shown).

## Discussion

In our cohort of PrEP-using MSM, we found the COVID-19 pandemic has impacted access to and utilization of numerous sexual health services: a quarter of the cohort reported challenges while attempting to access PrEP, HIV testing, or STD testing. Despite PrEP prescription guidelines indicating quarterly HIV and STD testing for MSM at high risk for recurrent STIs,^9^ our study found that many participants had not received a STD test or HIV test in the previous 3 months. Moreover, the pandemic seems to have impacted PrEP use itself with a fifth of participants either indicating they discontinued PrEP use or changed how often they take PrEP because of COVID-19.

To contextualize these findings relating to delivery and utilization of services, we also examined changes in sexual behavior and found important variations by partner type and time period. Most participants indicated a decrease in sexual activity with casual partners, but fewer reported decreases with main partners. Examining longitudinal trends, there appeared to be a consistent pattern - the cohort’s mean level of sexual risk significantly dropped from February to April and then significantly increased from April to June. This extends the body of literature which has also documented reductions in sexual risk among MSM at the earlier stages of the pandemic^5^ and suggests that such reductions may be short-lived. However, there did not appear to be meaningful variation in the number of missed PrEP doses by calendar month among participants who remained on PrEP. Although evidence of period effects for PrEP adherence were not apparent, it is important to note that a fifth of the cohort reported discontinuing PrEP use altogether or taking PrEP less frequently. Further, as prescriptions typically need to be written every three months, it is possible that potential effects to PrEP adherence would not be captured due to our study’s time frame.

Our study has limitations. Given limited sample size, we were unable to test for demographic and geographic differences in our findings. Further because participants are from several states in the South, we were unable to link our findings to specific state-level policies to control COVID-19. All data are self-reported and are subject to social desirability biases. Findings may not be generalizable to other populations or settings.

Our findings underscore the need to maintain STD and HIV prevention efforts during the COVID-19 pandemic, as recent calls for action have stressed.^13,14^ Finding unique ways to deliver these essential services is needed. Guidance from the US Centers for Disease Control and Prevention and other sources offer a host of options to ensure appropriate care during the pandemic,^15,16^ including lab-only visits for indicated HIV/STD tests, at-home HIV/STD testing, self-testing for HIV via an oral swab-based test, extending the prescription of PrEP to a 90-day supply to decrease trips to the pharmacy, utilizing telemedicine when possible, and finally referring patients elsewhere if needed services cannot be provided. Moreover, a critical component of PrEP check-up visits is risk reduction counselling and incorporating discussions of COVID-19 risk into these discussions may be needed. For all sexual activities examined except kissing, there was substantial variability in the perceived level of COVID-19 acquisition risk, suggesting a need for clearer risk reduction messaging in clinical contexts^17^—and beyond. Indeed, public health messaging on COVID-19 risk and sex has largely been absent, although some local health departments have released guidance on safer sex during the pandemic.^18^

Continued monitoring of PrEP use, utilization of sexual health services, and sexual behaviors during the pandemic among PrEP-using MSM and other subpopulations is needed, including longitudinal studies powered to examine short- and long-term changes, qualitative investigations detailing specific barriers and facilitators to care, and analyses of pharmaceutical and medical claims data elucidating temporal changes in service utilization. Given our findings which suggest reduced access to and utilization of STD and HIV prevention services coupled with a continuation of behaviors which confer STD/HIV risk, ensuring appropriate delivery of these essential services and relevant risk-reduction messaging as this pandemic continues to unfold is critical.

## Data Availability

Data from this study are not publicly available.

## Disclosure

The authors have no conflicts of interest to disclose.

An abstract presenting findings from this study was accepted for the 2020 STD Prevention conference (Online conference, September 14-24, 2020).

